# Quantitative T1 and Effective Proton Density (PD*) mapping in children and adults at 7T from an MP2RAGE sequence optimised for uniform T1-weighted (UNI) and FLuid And White matter Suppression (FLAWS) contrasts

**DOI:** 10.1101/2024.06.28.24307535

**Authors:** Ayşe Sıla Dokumacı, Katy Vecchiato, Raphael Tomi-Tricot, Michael Eyre, Philippa Bridgen, Pierluigi Di Cio, Chiara Casella, Tobias C. Wood, Jan Sedlacik, Tom Wilkinson, Sharon L. Giles, Joseph V. Hajnal, Jonathan O’Muircheartaigh, Shaihan J. Malik, David W. Carmichael

**Affiliations:** Biomedical Engineering Department, School of Biomedical Engineering and Imaging Sciences, King’s College London, London, United Kingdom; London Collaborative Ultra high field System (LoCUS), London, United Kingdom; Department of Forensic and Neurodevelopmental Sciences, Institute of Psychiatry, Psychology and Neuroscience, King’s College London, London, United Kingdom; Centre for the Developing Brain, School of Biomedical Engineering and Imaging Sciences, King’s College London, London, United Kingdom; MR Research Collaborations, Siemens Healthcare Limited, Frimley, United Kingdom; Children’s Neurosciences, Evelina London Children’s Hospital at Guy’s and St Thomas’ NHS Foundation Trust, London, UK; Department of Neuroimaging, Institute of Psychiatry, Psychology and Neuroscience, King’s College London, London, United Kingdom; Robert Steiner MR Unit, Medical Research Council Laboratory of Medical Sciences, Hammersmith Hospital Campus, Du Cane Road, London, UK; Mansfield Centre for Innovation, Imaging Sciences, Institute of Clinical Sciences, Imperial College London, Hammersmith Hospital Campus, Du Cane Road, London, UK; Guy’s and St Thomas’ NHS Foundation Trust, London, United Kingdom; MRC Centre for Neurodevelopmental Disorders, King’s College London, London, United Kingdom

**Keywords:** MP2RAGE, Quantitative MRI, 7T, Ultra-High Field, T_1_ Mapping, PD Mapping

## Abstract

**Introduction:** Quantitative MRI is important for non-invasive tissue characterisation. In previous work we developed a clinically feasible multi-contrast protocol for T_1_-weighted imaging based on the MP2RAGE sequence that was optimised for both children and adults. It was demonstrated that a range of Fluid And White Matter Suppression (FLAWS) related contrasts could be produced while maintaining T_1_-weighted uniform image (UNI) quality, a challenge at higher field strengths. Here we introduce an approach to use these images to calculate effective proton density (PD*) and quantitative T_1_ relaxation maps especially for shorter repetition times (TR_MP2RAGE_) than those typically used previously.

**Methods:** T_1_ and PD* were estimated from the analytical equations of the MP2RAGE signal derived for partial Fourier acquisitions. The sensitivity of the fitting results was evaluated with respect to the TR_MP2RAGE_ and B_1_^+^ effects on both excitation flip angles and inversion efficiency and compared to vendor T_1_ maps which do not use B_1_^+^ information. Data acquired for a range of individuals (aged 10-54 years) at the shortest TR_MP2RAGE_ (4000ms) were compared across white matter (WM), cortical grey matter, and deep grey matter regions.

**Results:** The T_1_ values were insensitive to the choice of different TR_MP2RAGE_. The results were similar to the vendor T_1_ maps if the B_1_^+^ effects on the excitation flip angle and inversion efficiency were not included in the fits. T_1_ values varied over development into adulthood, especially for the deep grey matter regions whereas only a very small difference was observed for WM T_1_. Effective PD maps were produced which did not show a significant difference between children and adults for the age range included.

**Conclusion:** We produced PD* maps and improved the accuracy of T_1_ maps from an MP2RAGE protocol that is optimised for UNI and FLAWS-related contrasts in a single scan at 7T by incorporating the excitation flip angle and inversion efficiency related effects of B_1_^+^ in the fitting. This multi-parametric protocol made it possible to acquire high resolution images (0.65mm iso) in children and adults within a clinically feasible duration (7:18 min:s). The combination of analytical equations utilizing B_1_^+^ maps led to T_1_ fits that were consistent at different TR_MP2RAGE_ values. Average WM T_1_ values of adults and children were very similar (1092ms vs 1117ms) while expected reductions in T_1_ with age were found for GM especially for deep GM.

## 1 Introduction

Quantitative MRI has an important role in the measurement of tissue microstructure and its alteration as part of healthy development and ageing (Deoni, 2010; Eminian et al., 2018; Lutti et al., 2014) and in a wide range of pathology (Adler et al., 2017; McDowell et al., 2018).

Many different approaches exist for longitudinal relaxation constant (T_1_) mapping (A. G. Teixeira et al., 2020; Brix et al., 1990; Christensen et al., 1974; Crawley & Henkelman, 1988; Deichmann & Haase, 1992; Deoni, 2007; Deoni et al., 2003; Frahm et al., 1986; Fram et al., 1987; Graumann et al., 1986; Gupta, 1977; Hahn, 1950; Helms et al., 2008; Henderson et al., 1999; Homer & Beevers, 1985; Leipert & Marquardt, 1976; Look & Locker, 1968, 1970; Lutti et al., 2014; Ma et al., 2023; Wright et al., 2008); however, a fast and widely used approach at 7 Tesla (T) for human brain is the Magnetization Prepared 2 Rapid Gradient Echoes (MP2RAGE) (Marques et al., 2010) sequence. The MP2RAGE sequence (Marques et al., 2010), which is an extension of the Magnetization Prepared RApid Gradient Echo (MPRAGE) sequence (Marques et al., 2010; Mugler & Brookeman, 1990; Van de Moortele et al., 2009), has been used at 7T to produce 3D structural T_1_-weighted brain images and T_1_ maps. This sequence acquires two Gradient Echo (GRE) images (INV1 and INV2) at two different inversion times (TI_1_ and TI_2_) following a non-selective adiabatic inversion pulse where the time between two consecutive inversion pulses is called TR_MP2RAGE_(Marques et al., 2010). These two images are combined to form the uniform T_1_-weighted image (UNI) which is insensitive to receive field (B_1-_) variability, proton density (PD), and T_2_* effects (Marques et al., 2010; Van de Moortele et al., 2009). T_1_ maps are typically produced using the UNI signal from look up tables (LUTs) which assume uniform B_1_^+^(Marques et al., 2010). Nonetheless, derived images and quantitative maps are still sensitive to transmit field (B1^+^) inhomogeneity effects (Marques et al., 2010).

Even though the MP2RAGE scan parameters can be optimised to reduce the B1^+^ effects, some residual B_1_^+^ bias is likely to be present in the T_1_ maps (Marques & Gruetter, 2013; Van de Moortele et al., 2009). To overcome this problem, Marques et al. (Marques & Gruetter, 2013) employed the MP2RAGE sequence(Marques et al., 2010) and SA2RAGE sequence (Eggenschwiler et al., 2012) that is used to calculate B1^+^ values assuming a single T_1_ value over the whole brain together (Marques & Gruetter, 2013). Two 2D look up tables relating either the T_1_ value to the MP2RAGE signal intensity and the B_1_^+^ value or the B_1_^+^ value to the MP2RAGE signal intensity and the T_1_ value were utilized iteratively to estimate both T_1_ and B_1_^+^(Marques & Gruetter, 2013). The B_1_^+^ effect was considered only for the excitation pulses and not for the adiabatic inversion pulse (Marques & Gruetter, 2013).

A study investigating the inter-site variability compared two datasets one of which was acquired with a low B_1_^+^-sensitive protocol like the one by Marques and Gruetter (Marques & Gruetter, 2013) while using a parallel transmit head coil for improved B_1_^+^ homogeneity (Haast et al., 2021). This demonstrated that B_1_^+^ inhomogeneity affects both the T_1_ values (Marques & Gruetter, 2013) and segmentation and cortical surface calculation (Haast et al., 2018), with differing values with and without B_1_^+^ inhomogeneity correction (Haast et al., 2021).

The MP2RAGE sequence can also be optimised to produce GM-dominant FLuid And White Matter Suppression (FLAWS) images (Beaumont et al., 2019, 2021; Martin et al., 2023; Massire et al., 2021; Müller et al., 2022; Tanner et al., 2012; Urushibata et al., 2019), which can be beneficial for deep GM visualisation and potentially improve lesion detection in multiple sclerosis and focal cortical dysplasia (Chen et al., 2018; Martin et al., 2023; Massire et al., 2021; Müller et al., 2022). It would be advantageous to be able to obtain T_1_ maps from these protocols, but T_1_ estimation is challenging due to the low grey matter (GM) signal in the bias-reduced UNI image which is used to calculate T_1_ unless additional bias-reduced FLAWS-related images such as the FLAWS_hc_ is used for this purpose (Beaumont et al., 2021).

A further clinically relevant quantitative value is proton density (PD) which is defined as the number of MR visible hydrogen protons in tissue (Mezer et al., 2016; Tofts, 2003). PD values have been shown to be altered in multiple sclerosis (Engström et al., 2014; Gracien et al., 2017), brain oedema in hepatic encephalopathy (Shah et al., 2008), and peritumoral oedema in malignant gliomas (Blystad et al., 2017). Quantitative PD maps can be obtained from MP2RAGE protocols; however, this has not been performed to the best of our knowledge.

A number of studies have proposed to obtain T_1_ maps by adding an additional GRE block to the MP2RAGE sequence (Marques et al., 2010) named as MP3RAGE: Hung et al. (Hung et al., 2013) determined T_1_ by fitting the inversion efficiency (*eff*) of the pulse as a free parameter if the pulse profile information was not available; Rioux et al. (Rioux et al., 2014) obtained T_1_ and flip angle (FA) information by using three different combinations of the images acquired at different inversion times using a LUT as in Marques et al. (Marques et al., 2010); and Olsson et al. (Olsson et al., 2022) fitted T_1_ and B_1_^+^ simultaneously by using closed-form approximations of the signal equations for small FAs while assuming an *eff* value of 0.96.

In this study, we aimed to determine if high quality T_1_ and effective PD (PD*) maps could be generated from a 7T protocol with shorter than conventional repetition times (TR_MP2RAGE_), previously optimised to generate UNI and FLAWS-related contrasts in a single scan (Dokumacı et al., 2023). The term effective PD has been used because the T_2_* effects have not been considered(Cercignani et al., 2018; Weiskopf et al., 2013). To achieve this aim, analytical equations that account for partial Fourier acquisitions were derived and fitting was performed using the analytical equations in combination with B_1_^+^ maps while investigating the effects of B_1_^+^ both on the excitation flip angles and the inversion efficiency (*eff)*. Results using protocols with different TR_MP2RAGE_ values were compared. Data from children and adults using the short TR_MPRAGE_ (=4000ms) were acquired and changes in T_1_ with age were investigated.

## 2 Methods

### 2.1 Theory

The time between two consecutive inversion pulses in an MP2RAGE sequence, TR_MP2RAGE_, includes two GRE blocks, the delay (TA) following the first inversion pulse until the start of the first GRE block, the delay (TB) between the two GRE blocks, and the remaining time (TC) following the second GRE block until the application of the next inversion pulse (Marques et al., 2010). Following the approach of Marques et al.(Marques et al., 2010), the MP2RAGE steady-state signal for the longitudinal magnetization and the modified signal equations accounting for the partial Fourier acquisition for the GRE blocks were derived as(Dokumaci AS, 2022):

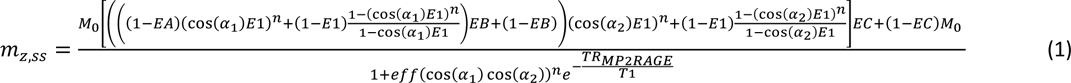

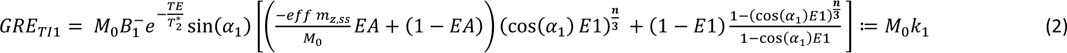

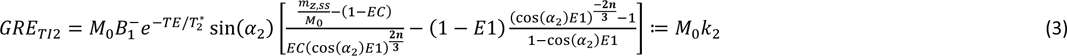

In Eqn. (1), EA = exp(-TA/T_1_), EB = exp(-TB/T_1_), EC = exp(-TC/T_1_), and E1 = exp(-TR_GRE_/T_1_). TR_GRE_ is the time between the small flip angle excitations in the GRE blocks (n in total). The efficiency of the inversion pulse is denoted as *eff* and defined as (Marques et al., 2010) 𝑀_𝑧,𝑖𝑛𝑣_ = −𝑒𝑓𝑓𝑀_𝑧_(0) where M_z,inv_ is the inverted longitudinal magnetisation and M_z_(0) is the initial longitudinal magnetisation. Equations (2) and (3) account for the partial Fourier factor of 6/8 in the first phase encoding (partition) direction, acquired in the innermost k-space loop. This results in a shift of the k-space centre from the middle of the n phase encoding blocks to (n/3 + 1) in the sequence implementation.

To fit T_1_ and M_0_ (PD) a nonlinear least-squares solver (*lsqnonlin*) in MATLAB (R2022b, The Mathworks, Natick, Massachusetts) was used to minimise the cost function:

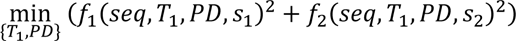 for f_1_ = s_1_ - PD*k_1_ and f_2_ = s_2_ - PD*k_2_ where s_1_ and s_2_ are the measured GRE_TI1_ and GRE_TI2_ signals, respectively with the theoretical values of PD*k_1_ and PD*k_2_ from Eqns. (2) and (3). The term *seq* represents the imaging parameters TR_GRE_, TI_1_, TI_2_, TR_MP2RAGE_, n, α_1_, α_2_ whose values are known. The terms k_1_ and k_2_ are not functions of PD but depend on T_1_ and the sequence parameters.

The fitting “options” for the lsqnonlin function were MaxIterations = 100000; MaxFunctionEvaluations = 400000; FunctionTolerance = 1e-20; FiniteDifferenceStepSize = 1e-2; and OptimalityTolerance = 1e-10. The T_1_ search range was 200ms to 30000ms while for the unscaled PD 0 to 100000 was used. The initial values for T_1_ and PD were 1500ms and 20000, respectively.

### 2.2 Different Fits for Calculating T_1_

To explore the weighting of the two likely factors contributing to spatial effects (inversion efficiency (*eff*) and B_1_^+^ variability) different fits were performed using the nonlinear least-squares algorithm:

**Fit 1** excluded B_1_^+^ information in the excitation flip angles (α_1_ and α_2_) and assumed *eff*=1 throughout the whole 3D image. **Fit 2** included B_1_^+^ information in the excitation flip angles (α_1_ and α_2_) but still assumed *eff* =1. Lastly, **Fit 3** included both the B_1_^+^ information and variable *eff*.

For all fits, the phase information was used to determine the polarity of the magnetization due to T_1_ recovery at the first inversion time (TI_1_).

The effect of TR_MP2RAGE_ on the fits was investigated. In addition, T_1_ maps from the vendor which are determined from a LUT without using B_1_^+^ information (Marques et al., 2010) were included for comparison. The function *colorbarpzn* (He, 2024) running in MATLAB was used to create the blue-white-red colormaps and colorbars.

### 2.3 Data Acquisition

All data was obtained under institutional ethical approval (REMAS 8700 and 18/LO/1766) and following informed consent. For all scans, a MAGNETOM Terra 7T system (Siemens Healthcare, Erlangen, Germany) using a single transmit (1Tx/32Rx) coil (Nova Medical) was employed.

The summary of scan protocols is given in Table 1. Most of the data were acquired using the protocol optimized in our previous study (Dokumacı et al., 2023) that confers the benefits of short scan time, high resolution, and high-quality FLAWS and UNI contrast options from a single scan. Firstly, data were obtained with different TR_MP2RAGE_ values in a small group of 4 healthy adults (32±2 years, 1f) to assess the invariance of the T_1_ estimate to the TR_MP2RAGE_ (Table 1a). Secondly, data from 7 healthy children (12±2 years, 2f) and 2 more adults making it 6 adults in total (35±9 years, 1f) were obtained to determine if expected age-related T_1_ changes could be observed; TR_MP2RAGE_=4000ms was used for all the scans in Table 1b except for Protocol 3 which had a low sensitivity to B_1_^+^ with a longer TR_MP2RAGE_ (Marques et al., 2010) (TR_MP2RAGE_=8000ms).

**Table 1.**
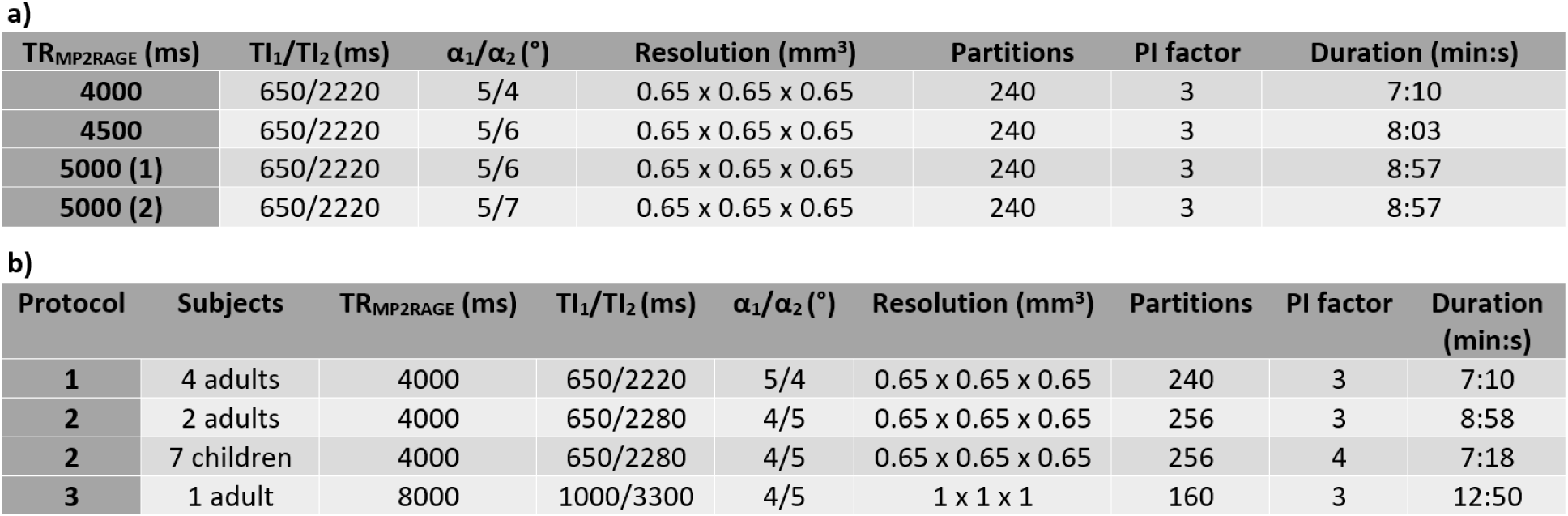
TI_1_/TI_2_ is the time between the inversion pulse and the centre of k-space for the 1^st^/2^nd^GRE block (Marques et al., 2010). Parallel imaging was employed using GRAPPA (Griswold et al., 2002). a) Scans obtained in 4 healthy adults: a partial Fourier factor of 6/8 was used in both phase encoding directions. b) Scan parameters applied in healthy children and adults. Protocol 3 had low sensitivity to B_1_^+^ but a longer TR_MP2RAGE_. A partial Fourier factor of 6/8 was used only in one direction (slice partial Fourier) for Protocols 2 and 3.

| $TR_{MP2RAGE}$ (ms) | $T_1/T_2$ (ms) | $\alpha_1/\alpha_2$ (°) | Resolution (mm<sup>3</sup>) | Partitions | PI factor | Duration (min:s) |
| --- | --- | --- | --- | --- | --- | --- |
| 4000 | 650/2220 | 5/4 | 0.65 x 0.65 x 0.65 | 240 | 3 | 7:10 |
| 4500 | 650/2220 | 5/6 | 0.65 x 0.65 x 0.65 | 240 | 3 | 8:03 |
| 5000 (1) | 650/2220 | 5/6 | 0.65 x 0.65 x 0.65 | 240 | 3 | 8:57 |
| 5000 (2) | 650/2220 | 5/7 | 0.65 x 0.65 x 0.65 | 240 | 3 | 8:57 |

b)
| Protocol | Subjects | $TR_{MP2RAGE}$ (ms) | $T_1/T_2$ (ms) | $\alpha_1/\alpha_2$ (°) | Resolution (mm<sup>3</sup>) | Partitions | PI factor | Duration (min:s) |
| --- | --- | --- | --- | --- | --- | --- | --- | --- |
| 1 | 4 adults | 4000 | 650/2220 | 5/4 | 0.65 x 0.65 x 0.65 | 240 | 3 | 7:10 |
| 2 | 2 adults | 4000 | 650/2280 | 4/5 | 0.65 x 0.65 x 0.65 | 256 | 3 | 8:58 |
| 2 | 7 children | 4000 | 650/2280 | 4/5 | 0.65 x 0.65 x 0.65 | 256 | 4 | 7:18 |
| 3 | 1 adult | 8000 | 1000/3300 | 4/5 | 1 x 1 x 1 | 160 | 3 | 12:50 |

To determine the effect of B_0_ inclusion on the *eff* of the inversion pulse, a B_0_ map (GRE sequence with 3 echoes TE_1_/TE_2_/TE_3_ = 1.02ms/2.26ms/4.08ms and TR=10ms) was acquired in one subject. In addition to the B_0_ map, MP2RAGE data using 2 different protocols (Protocols 2 and 3) and 2 different B_1_^+^ maps (sat_tfl and AFI (Yarnykh, 2007)) were obtained where sat_tfl refers to the B_1_^+^ method that uses a slice-selective preconditioning pulse in combination with a turbo FLASH readout (Chung et al., 2010; Fautz et al., 2008).

### 2.4 T_1_ and PD* estimation and analysis

Data preprocessing consisted of the following steps performed in MATLAB (R2022b, The Mathworks, Natick, Massachusetts): 1) using the FLAWS_hco_ images a brain mask was produced to limit the fit to brain and surrounding cerebrospinal fluid, 2) B_1_^+^ map processing, 3) B_1_^-^ bias field calculation for PD* map generation, and 4) determining the inversion efficiency (*eff*) maps which are explained in detail below.

#### 2.4.1 Producing the brain mask

FLAWS_hco_ images were generated using the INV1 and INV2 images (Beaumont et al., 2021; Dokumacı et al., 2023; Tanner et al., 2012) and converted to NIfTI using the *dicm2nii* function (Li et al., 2016). A brain mask was calculated from a segmentation performed on the FLAWS_hco_ image in SPM12 (Penny et al., 2007). Regions were assigned to be within the brain if they had probability of *p*>0.99 of being either white matter (WM), grey matter (GM), or cerebrospinal fluid (CSF). The resulting binary mask was edited to produce a closed mask with the application of the *imclose* and *imfill* morphological operators in MATLAB.

#### 2.4.2 B_1_^+^ map processing

B_1_^+^ maps were smoothed using the hMRI toolbox (Tabelow et al., 2019) and co-registered and resliced using the FLAWS_hco_ image (Beaumont et al., 2021; Dokumacı et al., 2023; Tanner et al., 2012) as the reference and the B_1_^+^ magnitude images as the source in SPM12 (Penny et al., 2007). The relative B_1_^+^ values for each voxel were multiplied by the nominal flip angles to define the actual flip angles subsequently used in the fitting process.

#### 2.4.3 B_1_^-^ related bias field calculation

Maps of the B_1_^-^ related signal bias in the PD* fits were produced using SPM12(Penny et al., 2007) using the following parameters in the *Segment* module: extremely light regularisation (0.00001), Bias FWHM of 30 mm cutoff, and 3 different tissue types were chosen because the fits were masked.

#### 2.4.4. Determining the inversion efficiency (eff) maps

The adiabatic inversion pulse was a hyperbolic secant (Baum et al., 1985; Silver et al., 1984) whose *eff* was determined using Bloch simulations for each spatial location for all subjects independently using the RF pulse information exported from the DICOM files in combination with the B_1_^+^ maps and, where available, B_0_ maps.

### 2.5 Data Analysis in FreeSurfer

To determine the T_1_ and PD* values for different brain regions, UNI images were segmented in FreeSurfer 7.3 (Fischl, 2012). For this purpose, the UNI images were masked with the same closed masks that were used in the fitting process. Masked images were denoised using the *DenoiseImage* function (Manjón et al., 2010) in Advanced Normalization Tools (ANTs)(Avants et al., 2009). A variant of the N3 Bias Correction algorithm(Sled et al., 1997, 1998), N4 Bias Correction algorithm (*N4BiasFieldCorrection* function) (Tustison et al., 2010) implemented in the ANTs (Avants et al., 2009) package was used with the default parameters (spline spacing b = 180; convergence c = [50x50x50x50, 0]). The masks were part of the inputs for both functions. Images were reoriented to standard orientation using the *fslreorient2std* function in FSL (FMRIB Software Library) (Jenkinson et al., 2012; Smith et al., 2004).

These images were segmented using the *recon-all* function (Collins et al., 1994; Dale et al., 1999; Fischl, 2004; Fischl et al., 2001, 2002; Fischl, Sereno, & Dale, 1999; Fischl, Sereno, Tootell, et al., 1999; Fischl & Dale, 2000; *Non-Uniform Intensity Correction*. http://Www.Bic.Mni.Mcgill.ca/Software/N3/Node6.Html, n.d.; Sled et al., 1997, 1998) of FreeSurfer(Fischl, 2012) with -all (to perform all including subcortical segmentations) and -cm (to conform to the maximum resolution) options. All segmentations were checked via visual assessment by using the Subcortical Automatic Segmentation (aseg) and Automatic Cortical Parcellation (aparc) files produced by FreeSurfer overlaid on original T_1_-weighted images. If the UNI image segmentation was not successful, the steps above were applied to the FLAWS_hco_ images to be used as the FreeSurfer input. In one subject, parameters used for all other subjects did not result in a good segmentation and therefore were adjusted - a spline spacing of b = 100 and convergence c = [1000, 0] were used to get an improved result.

The intersection of the FreeSurfer and SPM masks were used for WM while adding another constraint to include the values with T_1_ ≤ 2000 ms to avoid contamination from CSF due to imperfect segmentation; for cortex the additional constraint was T_1_ ≤ 2500 ms. For deep GM regions, only the FreeSurfer segmentations were used because of the high constraint in the SPM segmentation (*p*>0.99) leading to region of interests including very few voxels (in the extreme cases without any voxels) but, like the cortex, voxels with T_1_ ≤ 2500 ms were included in the masks. Regional T_1_ mean values and standard deviations (SD) were calculated for the group of subjects who had the TR_MP2RAGE_=4000ms protocols to see how the values within different brain regions change by age and to establish values in children at 7T.

## 3 Results

The pulse profile with the inversion efficiency simulated using the B_0_ and B_1_^+^ maps from the scan where Protocols 2 and 3 were compared are given in Figure 1. As seen in Figure 1a-b, the inversion pulse is very efficient for a large range of B_1_^+^ and off-resonance values. For this slice, the inversion efficiency mean value ± standard deviation was 0.995 ± 0.023 with a minimum value of 0.477. Without the inclusion of B_0_ information in the inversion efficiency calculations, the inversion efficiency for this slice was simulated as 0.995 ± 0.023 with a minimum of 0.478. Inversion efficiency over the 3D volume was calculated as 0.990 ± 0.048 and 0.990 ± 0.047 with and without the inclusion of the B_0_ map, respectively.

**Figure 1.**
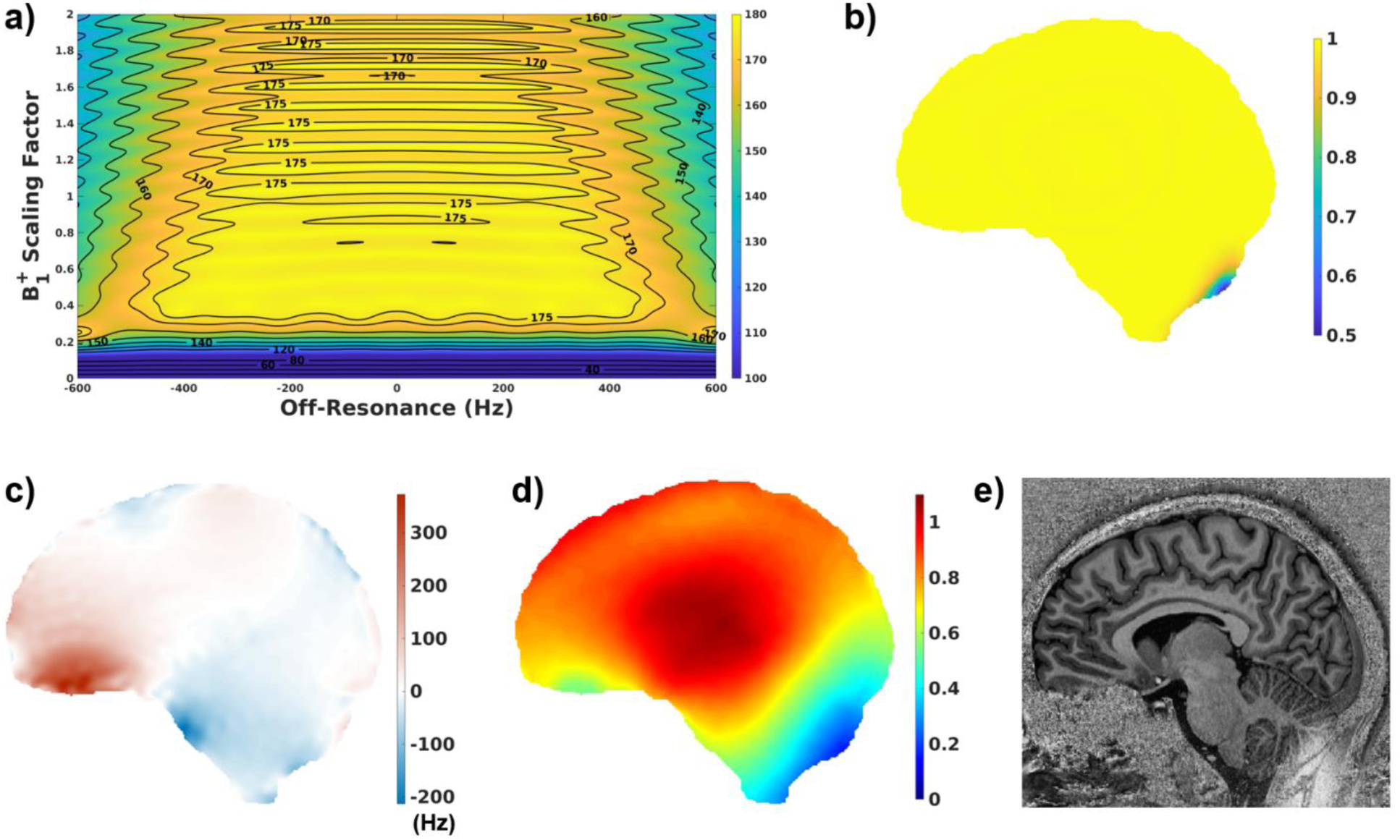
a) Pulse profile using a range of B_0_ and B_1_^+^ values for this pulse which is insensitive to a large range of B_0_ and B_1_^+^ values b) Inversion Efficiency (*eff*) map using c) B_0_ map from a slice acquired in this scan and d) the corresponding B_1_^+^ map (sat_tfl) e) the corresponding UNI image which shows a big change at the back of the cerebellum exactly where the *eff* deviates from 1.

The differences between the vendor T_1_ map which is based on a LUT that does not use B_1_^+^ information and the fit that uses B_1_^+^ information demonstrated the expected spatial effect in T_1_ due to the spatial variation in the B_1_^+^ field (Fig. 2). Different fits (Fits 1-3) were performed to investigate the contribution of the *eff* and B_1_^+^ variability to these spatial effects.

**Figure 2.**
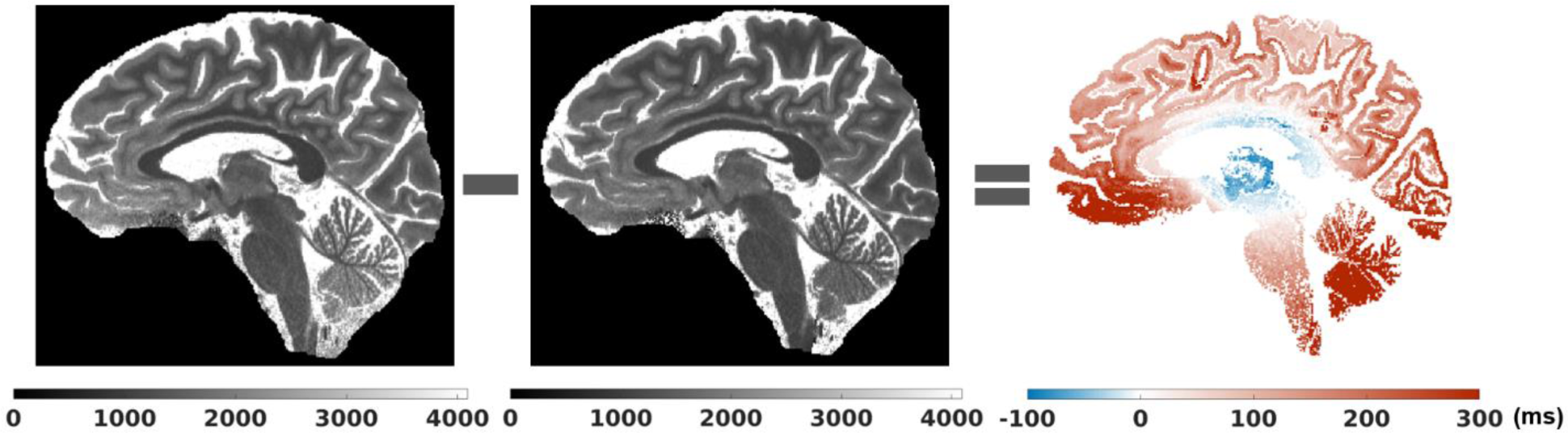
Vendor T_1_ map (a), the fit (b), and their difference for WM and GM (c) demonstrating the spatial effect related to B_1_^+^ non-uniformity.

Figure 3 shows the histograms of WM and cortex from the data of one adult subject using Protocol 1 for these different fits and the vendor T_1_ map. Supplementary Figure 1 shows the histograms from the deep GM regions. Table 2 lists the means and standard deviations for 4 adults including this subject (Adult 1) for WM and cortex.

**Figure 3.**
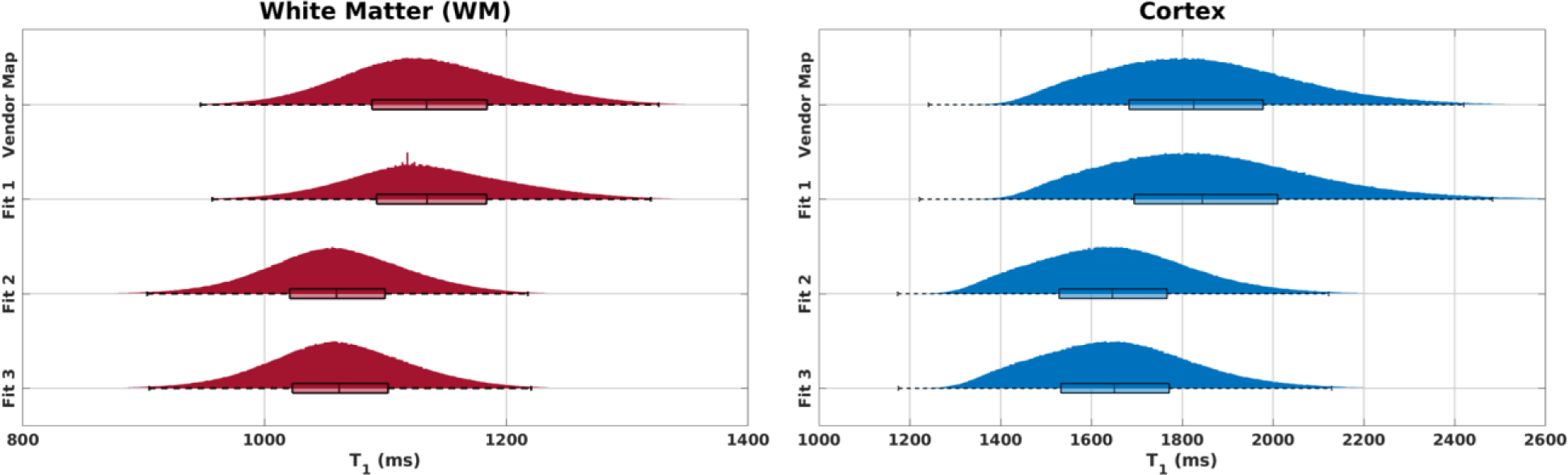
WM and cortex histograms for the vendor T_1_ map and different fits in one adult (Adult 1). Fit 1 does not consider the B_1_^+^ information like the vendor map. Fit 2 considers the B_1_^+^ information but assumes the inversion efficiency to be uniform (*eff* = 1) for each pixel. Fit 3 uses both the B_1_^+^ and *eff* maps for each pixel. Fit 1 produced the closest results to those of the vendor map. The results from Fits 2 and 3 were similar because of the high inversion efficiency of the adiabatic inversion pulse (*eff* = 0.994 ± 0.028 over all subjects).

**Table 2.** Means and SDs for WM and cortex T_1_ values (ms) using different fits for 4 adults (Protocol 1). As in Fig.3, the vendor map results are closest to those obtained with Fit 1 for all subjects. Likewise, Fits 2 and 3 provided very similar results due to the high inversion efficiency of the adiabatic pulse.

| $T_1$ Value (ms) | Adult 1 | | Adult 2 | | Adult 3 | | Adult 4 | |
| --- | --- | --- | --- | --- | --- | --- | --- | --- |
| Mean ± SD | WM | Cortex | WM | Cortex | WM | Cortex | WM | Cortex |
| Vendor Map | 1138 ± 72 | 1841 ± 217 | 1202 ± 81 | 1957 ± 229 | 1170 ± 67 | 1865 ± 227 | 1187 ± 77 | 1916 ± 218 |
| Fit 1 | 1139 ± 71 | 1866 ± 238 | 1201 ± 82 | 2021 ± 287 | 1168 ± 66 | 1901 ± 267 | 1184 ± 77 | 1968 ± 270 |
| Fit 2 | 1060 ± 62 | 1656 ± 178 | 1108 ± 67 | 1723 ± 184 | 1098 ± 54 | 1665 ± 191 | 1103 ± 60 | 1696 ± 177 |
| Fit 3 | 1063 ± 63 | 1660 ± 179 | 1111 ± 67 | 1729 ± 183 | 1100 ± 55 | 1670 ± 192 | 1105 ± 60 | 1702 ± 178 |

In all subjects, the mean and median values of the vendor map and Fit 1 were the closest as both ignored the B_1_^+^ information. For this highly efficient hyperbolic secant adiabatic inversion pulse with an *eff* of 0.994 ± 0.028 over 13 subjects, the results from Fit 2 and Fit 3 were very similar. Supplementary Table 1 shows the T_1_ results using different fits in deep GM regions where a similar trend is observed.

To investigate the effect of TR_MP2RAGE_ on T_1_ fits, 4 different TR_MP2RAGE_ protocols (Table 1a) were compared using the Fit 3 estimation (accounting for B_1_^+^ effects on both the excitation flip angles and the *eff*). In Figure 4, the T_1_ means and standard deviations for different brain regions with different TR_MP2RAGE_ values are shown. The standard deviation at each TR_MP2RAGE_ corresponds to the physiological variation of the T_1_ over the whole brain. The horizontal dotted lines correspond to the mean across TR_MP2RAGE_. The mean values and the uncertainty of the average using these different protocols are given above the lines. The uncertainty of the average is very small for each brain region in all adults which indicates the high precision of the T_1_ fits across TR_MP2RAGE_.

**Figure 4.**
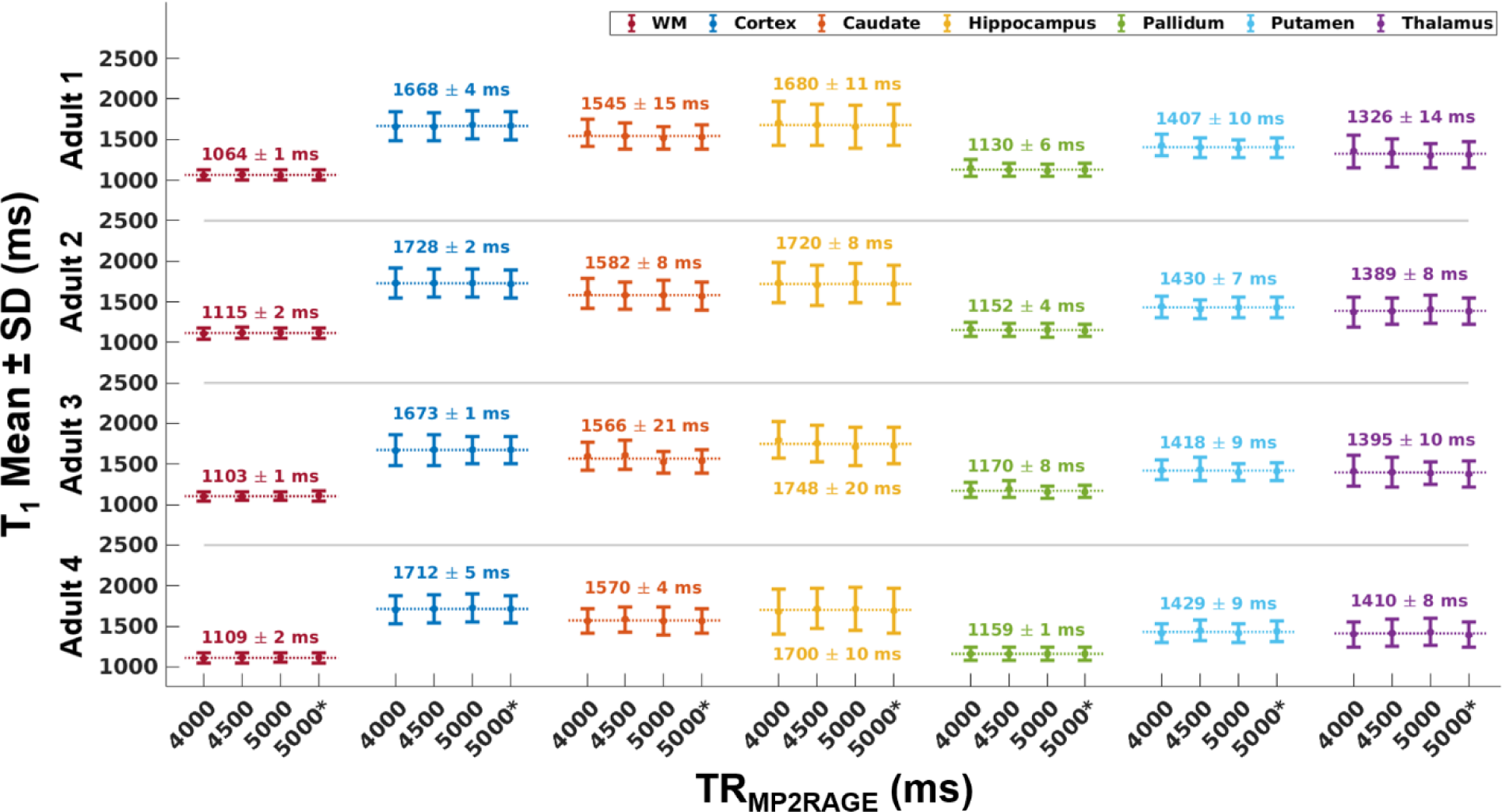
T_1_ measurements using Fit 3 and protocols with different TR_MP2RAGE_ values in 4 adults. The horizontal dotted lines correspond to the mean of the 4 measurements and the mean and uncertainty of the average is given above the line which indicates the small effect of these TR_MP2RAGE_ on the fits. TR_MP2RAGE_ 5000 and TR_MP2RAGE_ 5000* correspond to the protocols 5000 (1) and 5000 (2) in Table 1a, respectively.

After investigating the effect of TR_MP2RAGE_ on the fits, data acquired at TR_MP2RAGE_=4000ms were compared in children and adults. Figure 5 shows the results from different protocols (see Table 1b for protocol summaries) for WM, cortex and deep GM structures in children and adults. The difference between children and adult T_1_s was significant for GM regions especially for caudate and putamen. WM values were very similar for both populations.

**Figure 5.**
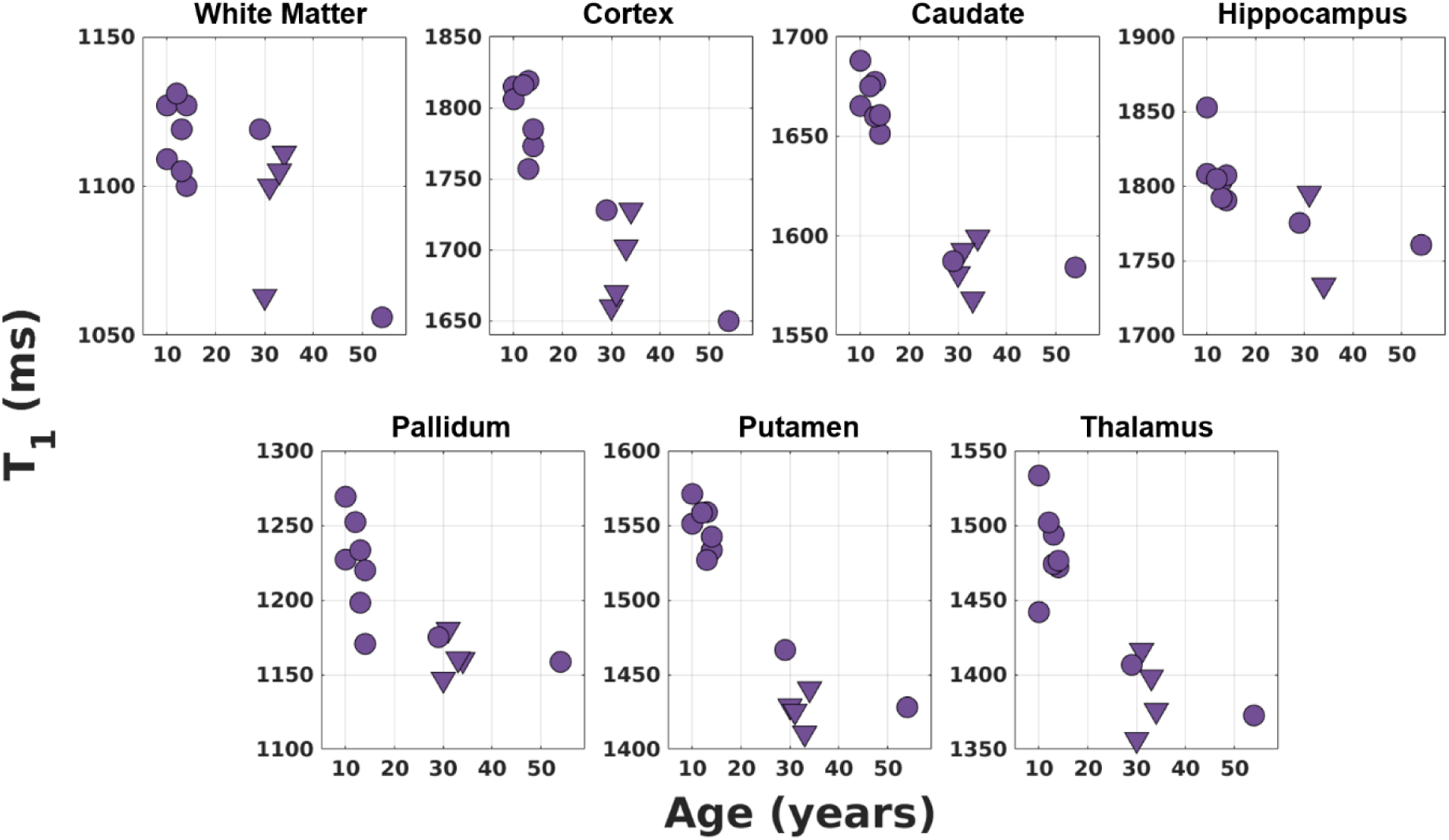
T_1_ fits (Fit 3) using different protocols (triangles correspond to Protocol 1 and disks correspond to Protocol 2 in Table 1b) with the same TR_MP2RAGE_ value (4000ms) in 7 children and 6 adults for different brain regions. The T_1_ values of the children are generally higher compared to those of the adults for the GM regions especially for caudate and putamen. WM values of the children (12 ± 2 years) and adults (35 ± 9 years) are very similar.

The plots for PD* values in different brain regions using different protocols in children and adults are shown in Figure 6. There is not a prominent difference between children and adult PD* values. Table 3 summarizes the means and standard deviations for T_1_ and PD* values of the children and adult populations.

**Figure 6.**
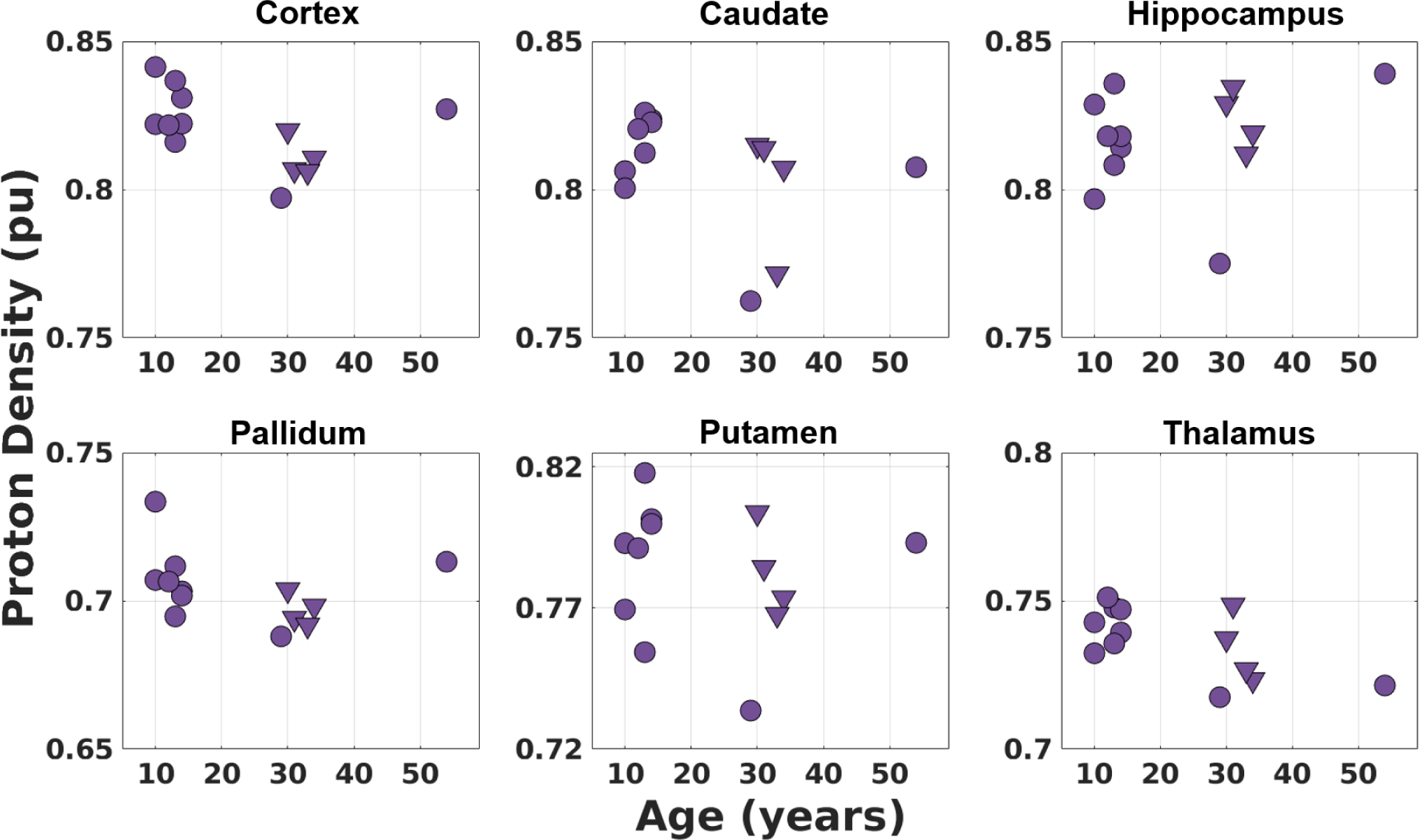
PD* values (Fit 3) using different protocols (triangles correspond to Protocol 1 and disks correspond to Protocol 2 in Table 1b) with the same TR_MP2RAGE_ value (4000ms) in 7 children and 6 adults for different brain regions. There is not a large difference between the children and adult PD* values although thalamus PD* values suggest a very slight decrease by age. The PD* values were scaled with respect to the WM mean which was assumed to be 0.69(Weiskopf et al., 2013). No T_2_* correction was made which would include a similar multiplication factor for WM and GM.

**Table 3.**
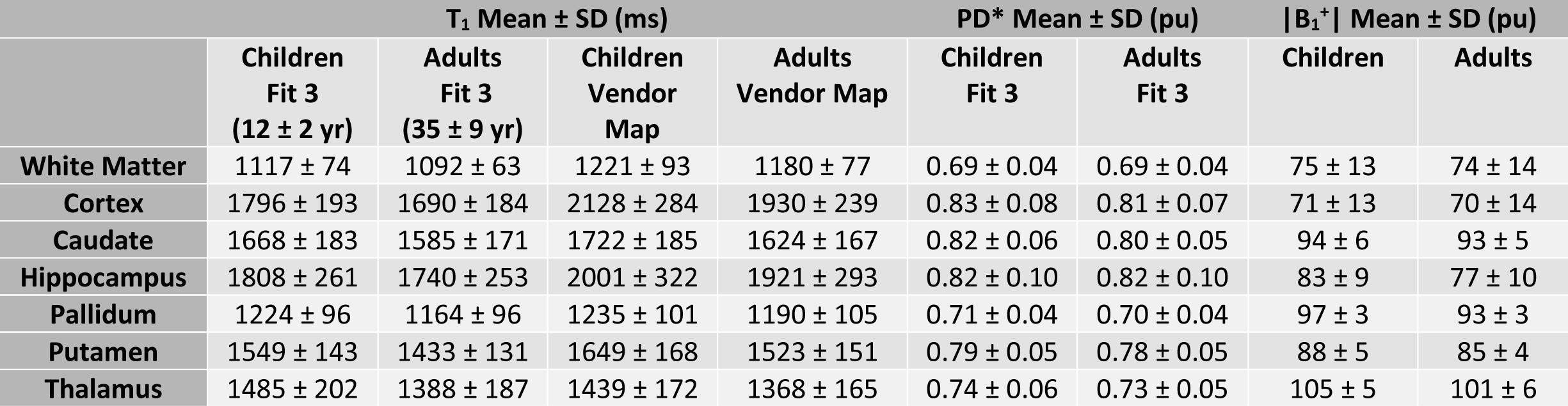
Means and standard deviations (SD) of T_1_, PD*, and B_1_^+^ in different brain regions of children and adults. The standard deviations reflect the physiological variation of tissue T_1_ across the brain. The difference between the T_1_ values of children and adults is significant for GM regions while the WM T_1_ values are very close. The PD* results of children and adults are very similar as well.

| | $T_1$ Mean ± SD (ms) | | | | PD* Mean ± SD (pu) | | $\vert B_1^+\vert $ Mean ± SD (pu) | |
| --- | --- | --- | --- | --- | --- | --- | --- | --- |
| | Children<br>Fit 3<br>(12 ± 2 yr) | Adults<br>Fit 3<br>(35 ± 9 yr) | Children<br>Vendor<br>Map | Adults<br>Vendor<br>Map | Children<br>Fit 3 | Adults<br>Fit 3 | Children | Adults |
| White Matter | 1117 ± 74 | 1092 ± 63 | 1221 ± 93 | 1180 ± 77 | 0.69 ± 0.04 | 0.69 ± 0.04 | 75 ± 13 | 74 ± 14 |
| Cortex | 1796 ± 193 | 1690 ± 184 | 2128 ± 284 | 1930 ± 239 | 0.83 ± 0.08 | 0.81 ± 0.07 | 71 ± 13 | 70 ± 14 |
| Caudate | 1668 ± 183 | 1585 ± 171 | 1722 ± 185 | 1624 ± 167 | 0.82 ± 0.06 | 0.80 ± 0.05 | 94 ± 6 | 93 ± 5 |
| Hippocampus | 1808 ± 261 | 1740 ± 253 | 2001 ± 322 | 1921 ± 293 | 0.82 ± 0.10 | 0.82 ± 0.10 | 83 ± 9 | 77 ± 10 |
| Pallidum | 1224 ± 96 | 1164 ± 96 | 1235 ± 101 | 1190 ± 105 | 0.71 ± 0.04 | 0.70 ± 0.04 | 97 ± 3 | 93 ± 3 |
| Putamen | 1549 ± 143 | 1433 ± 131 | 1649 ± 168 | 1523 ± 151 | 0.79 ± 0.05 | 0.78 ± 0.05 | 88 ± 5 | 85 ± 4 |
| Thalamus | 1485 ± 202 | 1388 ± 187 | 1439 ± 172 | 1368 ± 165 | 0.74 ± 0.06 | 0.73 ± 0.05 | 105 ± 5 | 101 ± 6 |

T_1_ values were fitted (Fit 3) over the 3D image using 2 different B_1_^+^ maps for Protocol 2 (TR_MP2RAGE_=4000ms) and it was also compared with Protocol 3 (TR_MP2RAGE_=8000ms) which is highly insensitive to B_1_^+^ changes. The B_1_^+^ sensitivities of all protocols used in this study are given in Supplementary Figure 2. The B_1_^+^ maps (sat_tfl and AFI) are shown in Supplementary Figure 3 for a slice with the corresponding T_1_ fits using Protocol 2.

Figure 7 demonstrates representative images and parameters maps that were produced using a single acquisition in under 7.5 minutes from a healthy child. The vendor T_1_ map and the T_1_ fit are displayed in the same range (0 to 4095 ms). The PD* map was scaled such that the WM mean would be equal to 0.69. No T_2_* correction was made for the PD* images. The T_2_* effect is expected to be similar for WM and GM.

**Figure 7.**
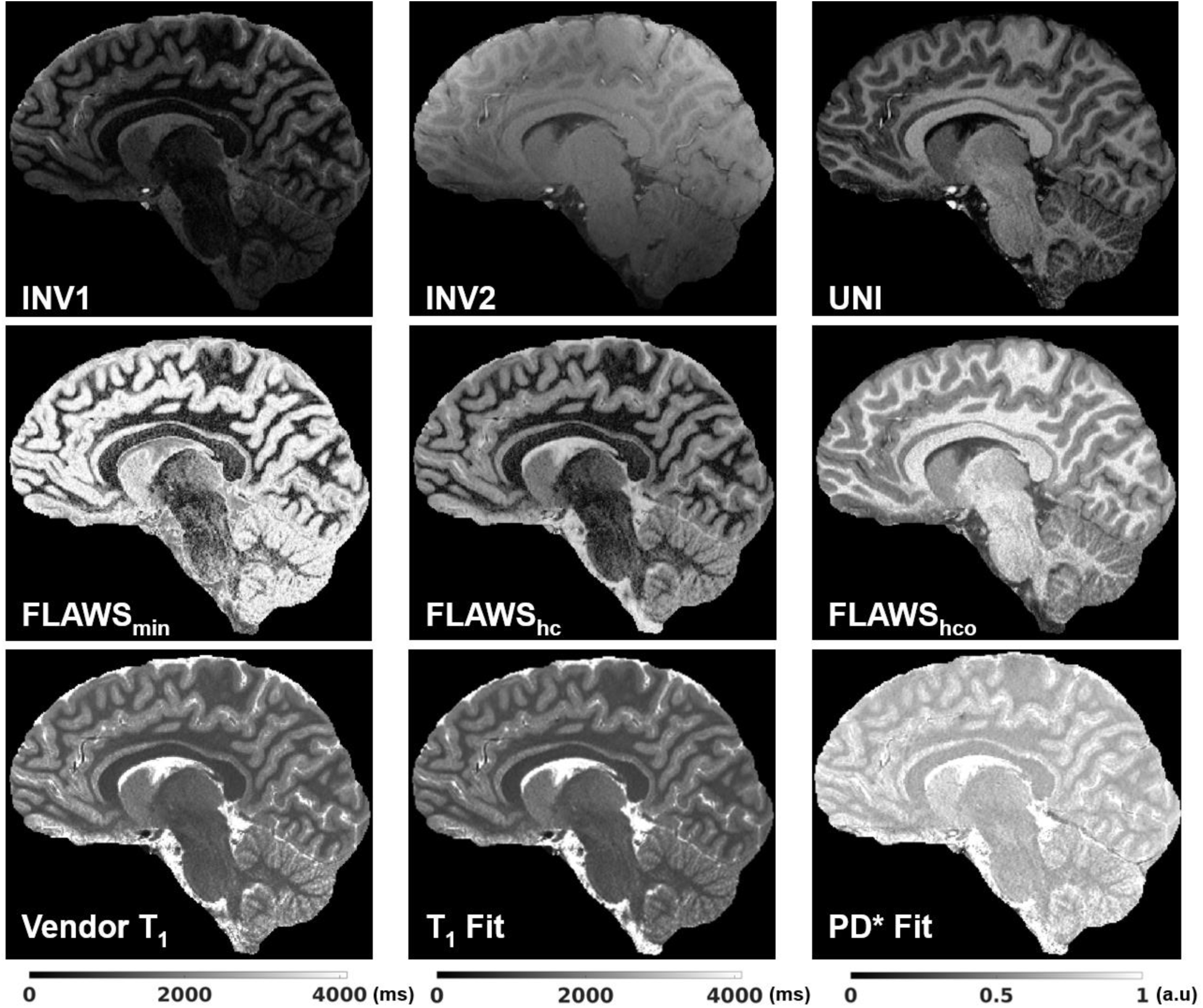
Different contrasts and parameter maps at 0.65 mm isotropic resolution were generated using a single MP2RAGE acquisition in 7:18 (min:s) from a healthy child. The vendor T_1_ map and the T_1_ fit are displayed in the same T_1_ range (0 to 4095 ms). The PD* image has not been corrected for T_2_* effects which would be a small factor due to the short TE (around 3 ms) and similar for WM and GM.

## 4 Discussion and Conclusion

In this study, we improved the accuracy of T_1_ maps and derived PD* images produced from our multi-contrast protocol that is optimised for UNI and FLAWS-related contrasts in a single scan (Dokumacı et al., 2023) at 7T by including B_1_^+^ effects for both the excitation flip angles and the inversion pulse in the fitting. This protocol made it possible to acquire high resolution images (0.65mm iso) in children and adults thanks to its short scan duration (7:18 min:s). T_1_ fitting results were consistent at different TR_MP2RAGE_ values (4000 ms, 4500 ms, and 5000 ms) for different brain regions (See Fig. 4). The fits resulted in a mean adult WM T_1_ value of 1092 ± 63 ms which was slightly lower than the values found in literature with the mean range of 1100-1400 ms (Marques et al., 2010; Marques & Norris, 2018; Wright et al., 2008). Mean T_1_ WM value for children (12 ±2 years) was 1117 ± 74 ms. This result being very similar to the adult T_1_ is not surprising as the myelination is relatively mature after 2-3 years of age with much smaller amplitude development into adulthood (Barkovich, 2000). Cortex and deep GM regions generally had slightly lower values compared to the literature adult T_1_ values (Caan et al., 2019) although they were similar and demonstrates the same pattern of lower T_1_ values in deep GM regions such as the thalamus. It was observed that the GM T_1_ values of children were higher than those of adults especially in deep GM regions such as caudate and putamen. This was consistent with the expected reductions in T_1_ with age from literature (Cho et al., 1997; Gracien et al., 2017). Cho and colleagues (Cho et al., 1997) demonstrated at 1.5T that T_1_ vs age relationship follows a quadratic curve with minimum T_1_ values observed for deep GM between ages of 38-48 while the inflection in the curve does not happen before 60 years for cortex. Figure 5 shows similar trends for deep GM and cortex with a higher T_1_ in the younger subjects. No significant difference was observed between the mean PD* values of children and adults (Figure 6 and Table 3). This is similar to the results from previous studies for this age range (Hagiwara et al., 2021; Saito et al., 2012). The PD* values in the thalamus (Figure 6) hints a very small decrease with age, but a larger study is required to confirm this result. Furthermore, it is possible that these small changes are related to small residual T_2_*-related changes in the PD* measure, despite the relatively short TE. A study by Callaghan and colleagues(Callaghan et al., 2014) found negative correlations between effective proton density and age in the putamen, pallidum, caudate nucleus, and the red nucleus with an echo time of 8.45ms at 3T.

It was confirmed that the B_1_^+^ has a large effect on the T_1_ values as found by Haast and colleagues (Haast et al., 2021) which necessitates future inclusion of the B_1_^+^ in T_1_ fits. This is particularly relevant where the protocol is sensitive to B_1_^+^ variations at shorter TR_MP2RAGE_ values which are a consequence of the requirements of short scan durations and high-resolution. The comparison of the B_1_^+^ corrected high-resolution images (0.65 mm isotropic) to lower-resolution images (1 mm isotropic) acquired with a low-B_1_^+^-sensitive protocol by Marques and Gruetter (Marques & Gruetter, 2013) revealed that by using any suitable B_1_^+^ map, the residual effects of B_1_^+^ variability on T_1_ maps can be removed. The accuracy of the B_1_^+^ map is important as it might affect the T_1_ results significantly for protocols that are more sensitive to B_1_^+^ variations. T_1_ values fitted using two different B_1_^+^ mapping methods in one subject were comparable (Table 4 and Supplementary Figure 3). It could be beneficial to use protocols that are less sensitive to B_1_^+^ variations while satisfying the short scan duration criterion for instance by combining CS methods (Candes & Wakin, 2008; Donoho, 2006; Lustig et al., 2007; Mussard et al., 2020; Puy et al., 2012; Trotier et al., 2019, 2022; Vasanawala et al., 2010) with longer TR_MP2RAGE_ values.

**Table 4.**
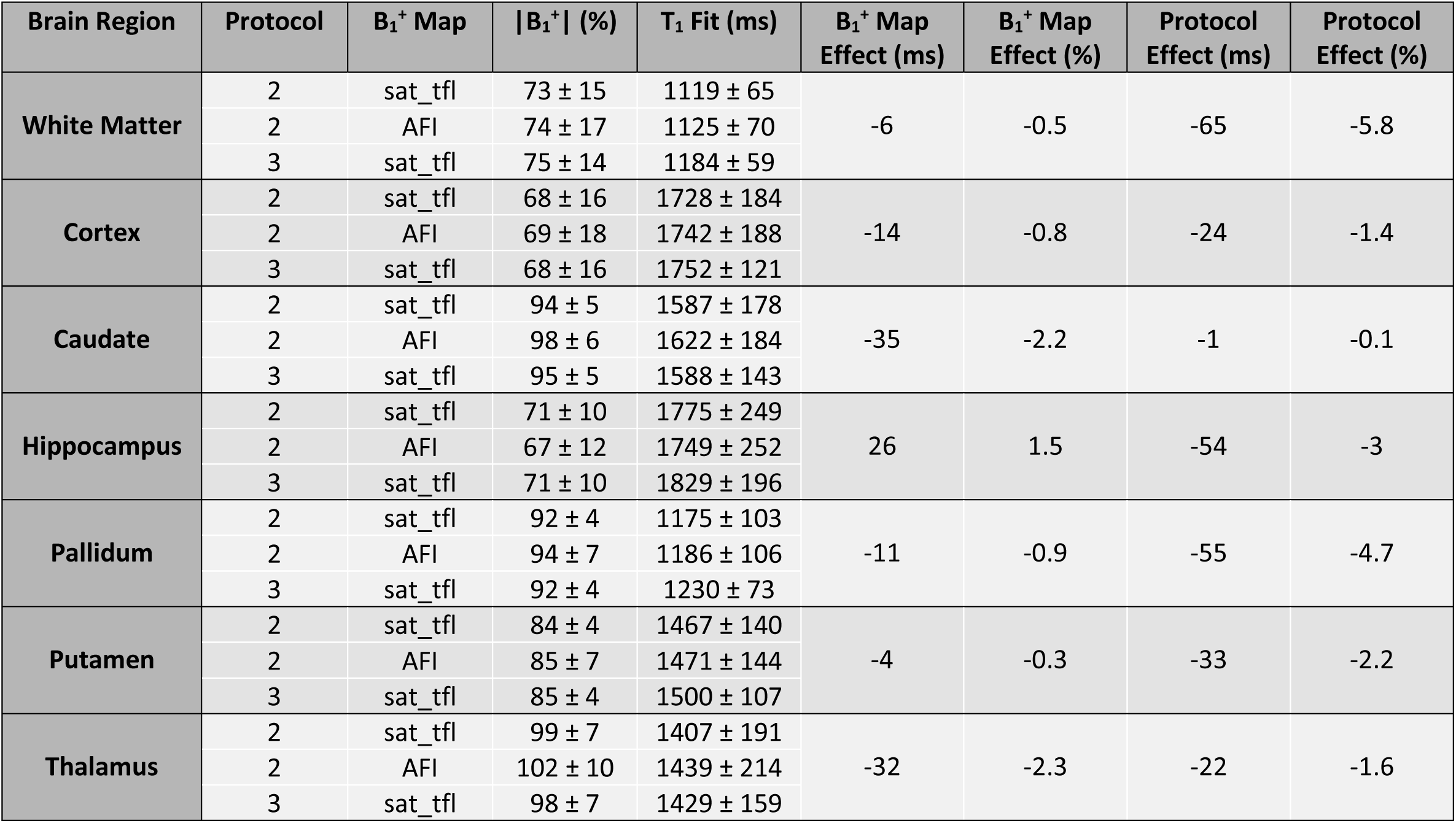
T_1_ value comparison (Fit 3) in the same subject using different B_1_^+^ maps (sat_tfl and AFI) and different protocols. Protocol 3 (TR_MP2RAGE_=8000ms) is less sensitive to the variations in B_1_^+^.

| Brain Region | Protocol | $B_1^+$ Map | $\vert B_1^+\vert $ (%) | $T_1$ Fit (ms) | $B_1^+$ Map Effect (ms) | $B_1^+$ Map Effect (%) | Protocol Effect (ms) | Protocol Effect (%) |
| --- | --- | --- | --- | --- | --- | --- | --- | --- |
| White Matter | 2 | sat_tfl | $73 \pm 15$ | $1119 \pm 65$ | -6 | -0.5 | -65 | -5.8 |
| | 2 | AFI | $74 \pm 17$ | $1125 \pm 70$ | | | | |
| | 3 | sat_tfl | $75 \pm 14$ | $1184 \pm 59$ | | | | |
| Cortex | 2 | sat_tfl | $68 \pm 16$ | $1728 \pm 184$ | -14 | -0.8 | -24 | -1.4 |
| | 2 | AFI | $69 \pm 18$ | $1742 \pm 188$ | | | | |
| | 3 | sat_tfl | $68 \pm 16$ | $1752 \pm 121$ | | | | |
| Caudate | 2 | sat_tfl | $94 \pm 5$ | $1587 \pm 178$ | -35 | -2.2 | -1 | -0.1 |
| | 2 | AFI | $98 \pm 6$ | $1622 \pm 184$ | | | | |
| | 3 | sat_tfl | $95 \pm 5$ | $1588 \pm 143$ | | | | |
| Hippocampus | 2 | sat_tfl | $71 \pm 10$ | $1775 \pm 249$ | 26 | 1.5 | -54 | -3 |
| | 2 | AFI | $67 \pm 12$ | $1749 \pm 252$ | | | | |
| | 3 | sat_tfl | $71 \pm 10$ | $1829 \pm 196$ | | | | |
| Pallidum | 2 | sat_tfl | $92 \pm 4$ | $1175 \pm 103$ | -11 | -0.9 | -55 | -4.7 |
| | 2 | AFI | $94 \pm 7$ | $1186 \pm 106$ | | | | |
| | 3 | sat_tfl | $92 \pm 4$ | $1230 \pm 73$ | | | | |
| Putamen | 2 | sat_tfl | $84 \pm 4$ | $1467 \pm 140$ | -4 | -0.3 | -33 | -2.2 |
| | 2 | AFI | $85 \pm 7$ | $1471 \pm 144$ | | | | |
| | 3 | sat_tfl | $85 \pm 4$ | $1500 \pm 107$ | | | | |
| Thalamus | 2 | sat_tfl | $99 \pm 7$ | $1407 \pm 191$ | -32 | -2.3 | -22 | -1.6 |
| | 2 | AFI | $102 \pm 10$ | $1439 \pm 214$ | | | | |
| | 3 | sat_tfl | $98 \pm 7$ | $1429 \pm 159$ | | | | |

Quantitative results are also affected by the accuracy of the segmentations. We used an additional precaution to prevent contamination by CSF voxels by limiting the masks for T_1_ values ≤ 2000 ms for WM and ≤ 2500 ms for GM. T1 restriction was found to have a negligible effect on all the values except for hippocampus which is an area more prone to segmentation errors (Supplementary Table 2).

One limitation of our study was that the B_0_ map was available for only one subject. However, due to the very high inversion efficiency of this hyperbolic secant adiabatic pulse (*eff* = 0.994 ± 0.028 over all subjects), the simulated B_0_ effect in the inversion efficiency using the map acquired in this subject was negligible. As seen in Figure 1, the efficiency deviates from 1 when B_1_^+^ is very low and it is not so related to B_0_. No separate correction was made for the exponential term^−𝑇𝐸/𝑇∗^ but TE was very short (about 3 ms). Another limitation was that the signal equations assumed the conventional mono-exponential decay (Labadie et al., 2014; Metere et al., 2017).

In conclusion, it was possible to produce quantitative T_1_ and PD* maps in children and adults at 7T in addition to the UNI and FLAWS-related contrasts from a single scan by using newly derived analytical equations required for partial Fourier acquisitions while incorporating the B_1_^+^ inhomogeneity effect both on the excitation flip angles and the inversion efficiency.

## Data Availability

The datasets that support the findings of this study are available upon request for any reasonable scientific purposes.

## Acknowledgments

The authors would like to acknowledge David Leitão, Ronald Mooiweer, and Oral Ersoy Dokumaci for valuable discussions.

This research was supported by GOSHCC Sparks Grant V4419, King’s Health Partners, in part by the Medical Research Council (UK) (grants MR/ K006355/1 and MR/LO11530/1) and Medical Research Council Center for Neurodevelopmental Disorders, King’s College London (MR/N026063/1), and by core funding from the Wellcome EPSRC Centre for Medical Engineering at King’s College London [WT203148/Z/16/Z]. J.O.M, K.V, and C.C were funded by a Sir Henry Dale Fellowship jointly by the Wellcome Trust and the Royal Society (206675/Z/17/Z). C.C was also funded by a grant from GOSHCC (VC1421). M.E was funded by Action Medical Research (GN2835) and the British Paediatric Neurology Association. This research was funded in whole, or in part, by the Wellcome Trust [WT203148/Z/16/Z and 206675/Z/17/Z] and by the National Institute for Health Research (NIHR) Biomedical Research Centre based at Guy’s and St Thomas’ NHS Foundation Trust and King’s College London and/or the NIHR Clinical Research Facility. The views expressed are those of the author(s) and not necessarily those of the NHS, the NIHR or the Department of Health and Social Care. For the purpose of open access, the author has applied a CC BY public copyright licence to any Author Accepted Manuscript version arising from this submission.

## Author Contributions

A.S.D: Conceptualization, Methodology, Software, Validation, Formal Analysis, Investigation, Resources, Data Curation, Writing – Original Draft, Writing – Review & Editing, Visualization.

K.V: Investigation, Resources, Data Curation, Writing – Review & Editing.

R.T-T: Methodology, Software, Data Curation, Writing – Review & Editing.

M.E: Software, Investigation, Resources, Data Curation, Writing – Review & Editing.

P.B: Investigation, Resources, Data Curation, Project Administration.

P.D.C: Investigation, Resources, Data Curation.

C.C: Investigation, Resources, Data Curation, Writing – Review & Editing.

T.C.W: Methodology, Software, Validation, Writing – Review & Editing.

J.S: Software, Data Curation, Writing – Review & Editing.

T.W: Software, Data Curation.

S.L.G: Resources, Writing – Review & Editing, Project Administration.

J.V.H: Methodology, Validation, Writing – Review & Editing, Visualization.

J.O.M: Methodology, Writing – Review & Editing, Visualization, Supervision, Project Administration, Funding Acquisition.

S.J.M: Conceptualization, Methodology, Software, Validation, Formal Analysis, Writing – Review & Editing, Visualization, Supervision, Funding Acquisition.

D.W.C: Conceptualization, Methodology, Validation, Formal Analysis, Investigation, Resources, Writing – Original Draft, Writing – Review & Editing, Visualization, Supervision, Project Administration, Funding Acquisition.

