## Supplementary Material for "Quantitative T1 and Effective Proton Density (PD*) mapping in children and adults at 7T from an MP2RAGE sequence optimised for uniform T1-weighted (UNI) and FLuid And White matter Suppression (FLAWS) contrasts"

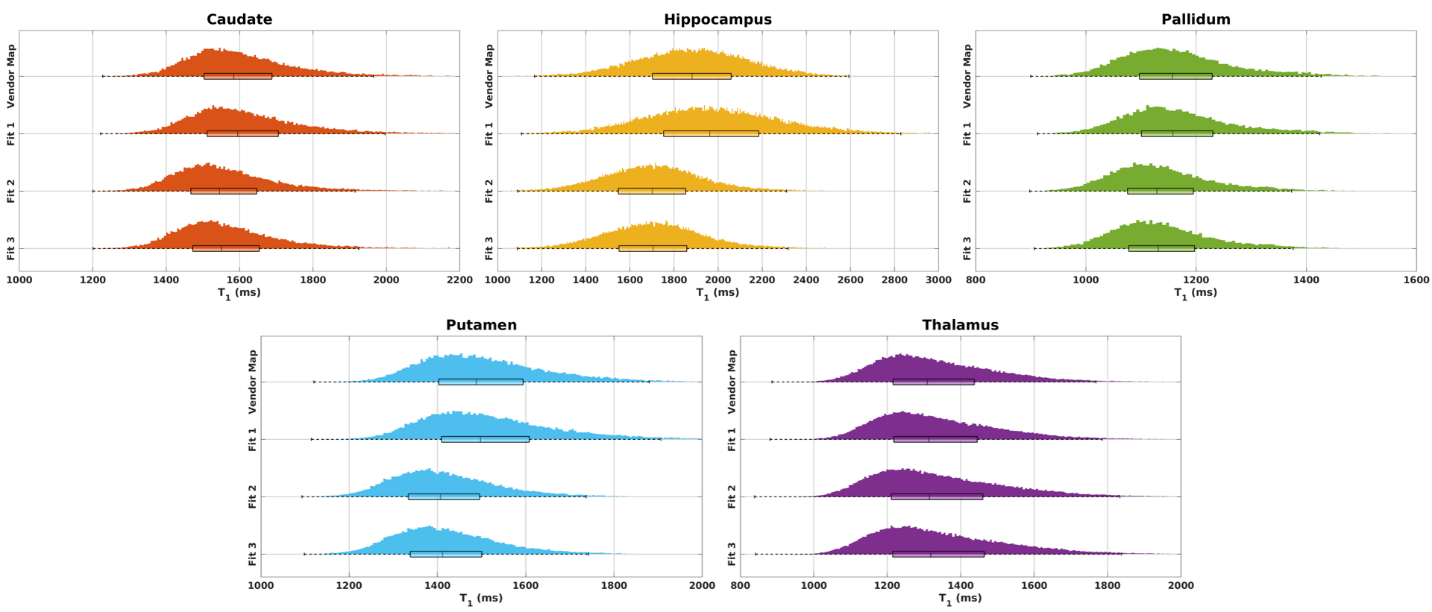
**Supplementary Material**

**Supplementary Figure 1.** T_1_ histograms of deep grey matter regions for the vendor T_1_ map and different fits in Adult 1. Fit 1 and the vendor map do not consider the B_1_^+^ information. Fits 2 and 3 both consider the B_1_^+^ information: Fit 2 assumes the *eff* to be 1 in each pixel whereas Fit 3 uses *eff* information for each pixel.

**Supplementary Table 1.** Means and SDs for deep grey matter T_1_ values (ms) using different fits for 4 adults (Protocol 1) showing the similar trend observed among different fits for WM and cortical T_1_ values.

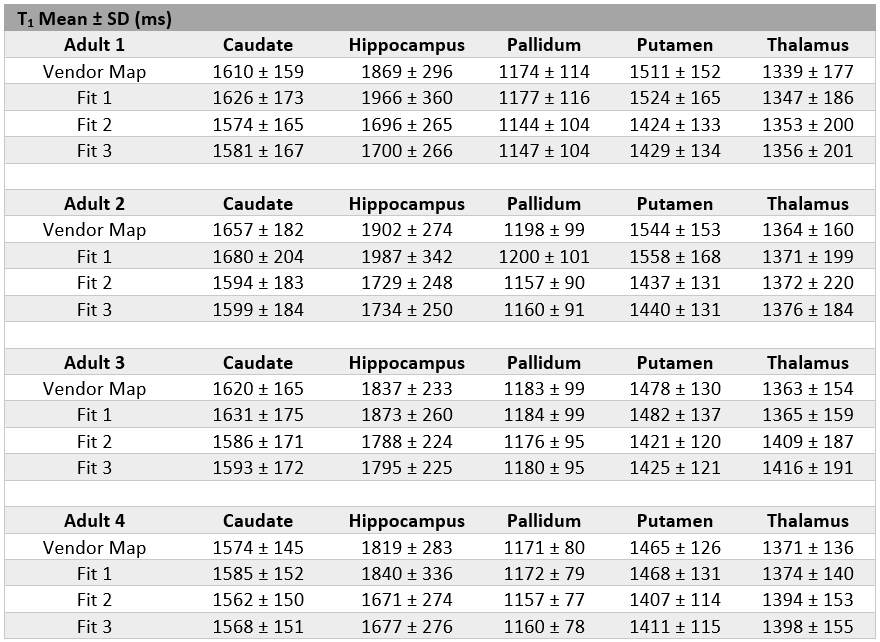

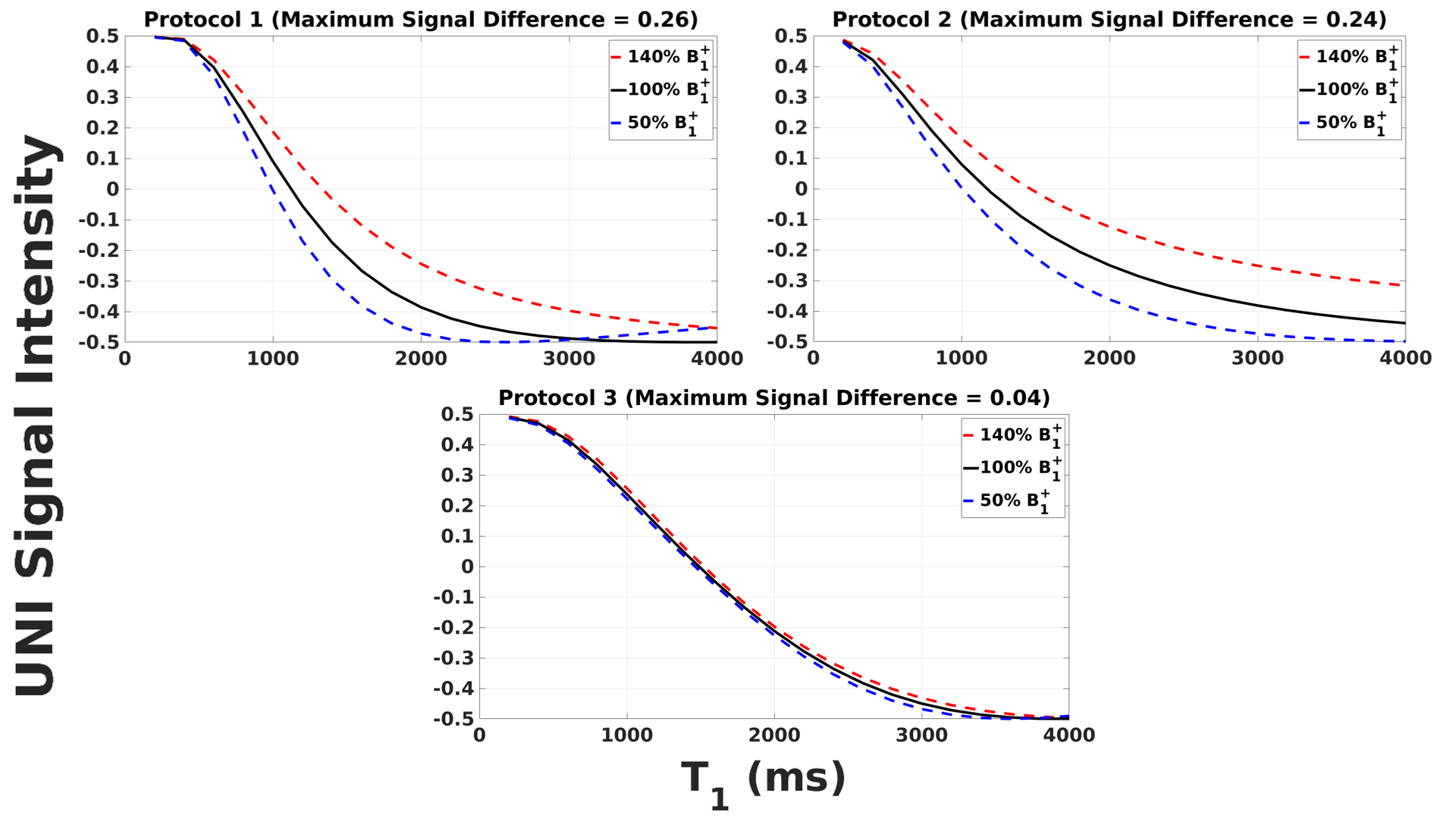
**Supplementary Figure 2.** B_1_^+^ sensitivity of different protocols used in this study. It is important to note that Protocol 3 has a TR_MP2RAGE_ value of 8000 ms whereas the other protocols have TR_MP2RAGE_ values of 4000 ms.

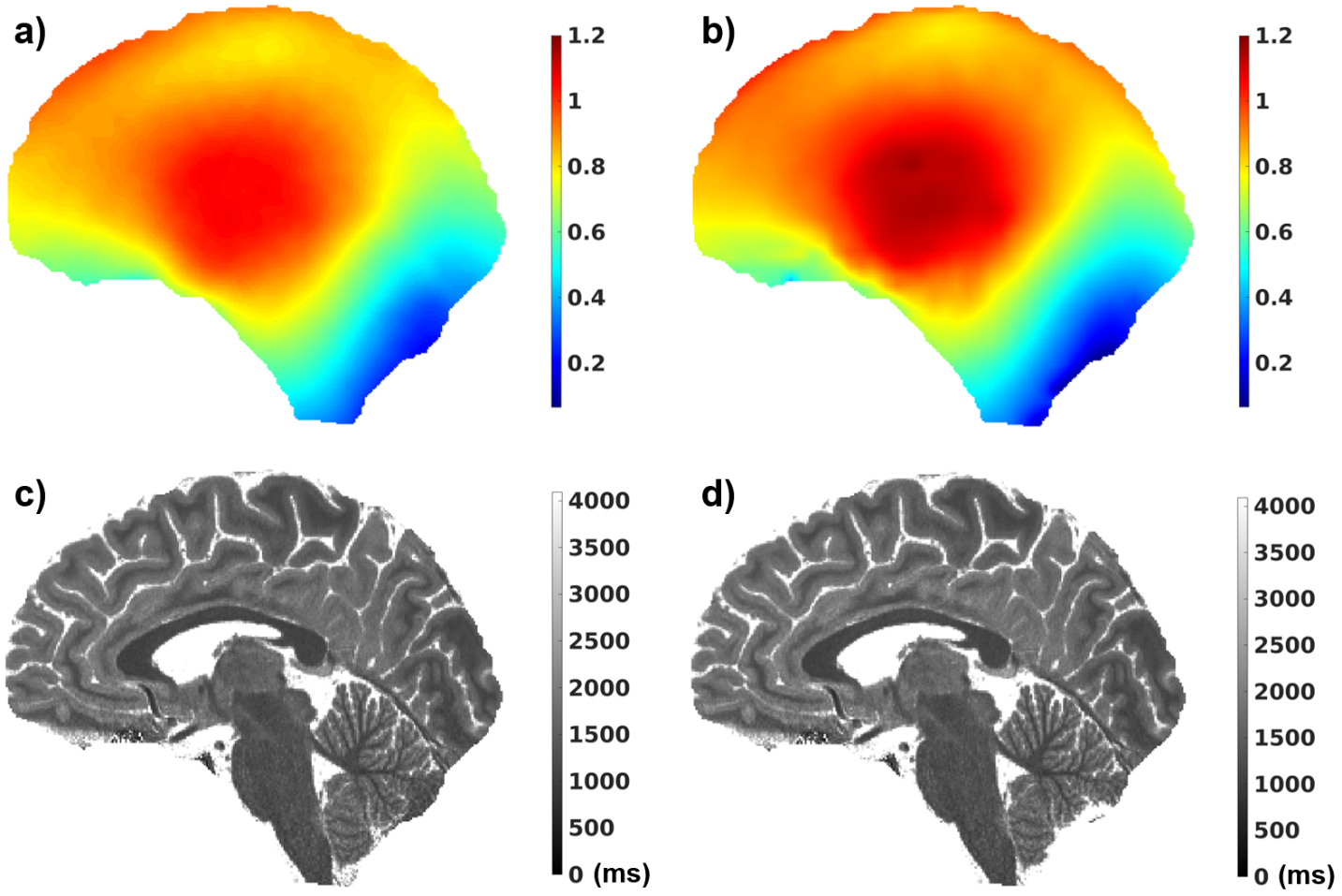

**Supplementary Figure 3.** Different B_1_^+^ maps applied in the same subject with the corresponding T_1_ fits. a) sat_tfl map b) AFI map c) T_1_ fit where sat_tfl was used to correct for B_1_^+^ d) T_1_ fit where AFI was used for B_1_^+^ correction.

**Supplementary Table 2.** Means and SDs of T_1_ values in children and adults for different brain regions (ms) using different masks. Intersection Mask refers to the common voxels between the FreeSurfer and SPM (*p*>0.99) segmentations. The SPM probability value was found to be too limited for deep GM regions sometimes resulting in an empty mask; therefore, only the FreeSurfer mask was used for deep GM. T_1_ restriction (≤ 2000 ms for WM and ≤ 2500 ms for GM) was not found to affect the results significantly except for hippocampus for which the CSF contamination was more prominent.

|  | **Intersection Mask &**  **T_1_ Restriction** | | **Intersection Mask &**  **No T_1_ Restriction** | | **FreeSurfer Mask &**  **T_1_ Restriction** | | **FreeSurfer Mask &**  **No T_1_ Restriction** | |
| --- | --- | --- | --- | --- | --- | --- | --- | --- |
|  | **Children** | **Adults** | **Children** | **Adults** | **Children** | **Adults** | **Children** | **Adults** |
| **WM** | 1117 ± 74 | 1092 ± 63 | 1118 ± 126 | 1093 ± 65 | 1177 ± 143 | 1142 ± 124 | 1185 ± 392 | 1145 ± 226 |
| **Cortex** | 1796 ± 193 | 1690 ± 184 | 1805 ± 366 | 1693 ± 302 | 1788 ± 225 | 1698 ± 229 | 1799 ± 568 | 1716 ± 551 |
| **Caudate** |  | |  | | 1668 ± 183 | 1585 ± 171 | 1680 ± 219 | 1593 ± 201 |
| **Hippocampus** |  |  |  |  | 1808 ± 261 | 1740 ± 253 | 1873 ± 560 | 1786 ± 497 |
| **Pallidum** |  |  |  |  | 1224 ± 96 | 1164 ± 96 | 1224 ± 97 | 1164 ± 102 |
| **Putamen** |  |  |  |  | 1549 ± 143 | 1433 ± 131 | 1550 ± 150 | 1434 ± 138 |
| **Thalamus** |  |  |  |  | 1485 ± 202 | 1388 ± 187 | 1491 ± 228 | 1394 ± 246 |
